# Resource conflicts leading to moral distress: A longitudinal study among physicians in Norway

**DOI:** 10.1101/2023.09.29.23295833

**Authors:** Ingrid Miljeteig, Reidun Førde, Karin Isaksson Rø, Fredrik Bååthe, Berit Bringedal

## Abstract

**Background:** The scarcity of resources represents ethical challenges and involves moral distress for health professionals. There are no longitudinal studies of moral distress among representative samples of physicians.

**Method:** Surveys of the Norwegian Physician Panel (NPP) compared the extent of moral distress in 2004 and 2021. Descriptive statistics and regression analysis were used in the study.

**Results:** Response rates were 67% (1004/1499) in 2004 and 70% (1639/2316) in 2021. That patient care is deprived due to time constraints is the most severe dimension of moral distress among physicians, and it has increased comparing 2021 with 2004 (68.3% in 2004 to 75.1% in 2021 reported “somewhat” or “very morally distressing”). Moral distress also increased concerning patients who “cry the loudest” get better and faster treatment than others. Moral distress was reduced on statements about long waiting times, treatment not provided due to economic limitations, deprioritisation of older patients, and acting against one’s conscience. Women reported higher moral distress than men in both years, and there were significant gender differences for six statements in 2021 and one in 2004. Though not consistently, the physicians’ age and workplace influenced the reported moral distress.

**Conclusion:** In both years, moral distress among physicians related to scarcity of or unfair distribution of resources was high. Moral distress associated with resource scarcity and acting against one’s conscience decreased, which might indicate improvements in the healthcare system. On the other hand, it might suggest that physicians have reduced their ideals or expectations or are morally fatigued.

## Background

Worldwide, healthcare providers face resource scarcity and complex priority-setting dilemmas. The COVID-19 pandemic enforced these dilemmas in most countries [1–4]. In the future, healthcare providers must likely handle more resource shortages due to an ageing population, increasing numbers of chronically ill patients and lack of qualified staff. Close attention should be paid to how the providers handle these challenges and their level of moral distress. After Jameton first introduced the concept of ‘moral distress’ in 1984, there has been an ongoing discussion on the definition [2]. Rushton is one among many who provide a more specified definition: “Moral distress ensues when clinicians recognise ethical conflicts and their responsibility to respond to them but are unable to translate their moral choices into ethically grounded action that preserves integrity” [5]. A high level of moral distress is correlated with reduced work performance and job satisfaction, a negative impact on the providers’ health, and an increased urge to resign [6–8]. Moral distress, related to having to give substandard care, has been studied in various ways, primarily among nurses, but also among other health care providers [9,10]. The results of these studies depend on how moral distress is defined, the measurement tools, and the study design [11–16].

In 2004, nine statements about work-related moral distress were presented to a representative sample of physicians in Norway (the Norwegian Physician Panel, NPP) [17]. The statements were drawn from a much-cited study by Kälvemark et al., who surveyed various health professionals [18]. The statements focused on experiences related to resource allocation, fairness concerns, and acting against one’s conscience. The Norwegian study showed that two-thirds of the physicians found time constraints and patient waiting times very or somewhat distressing. The physicians’ age, gender and working position (GP or hospital physicians) significantly impacted their responses. Other studies also show that being female, younger, and their speciality are predictors of higher moral distress among health care providers [19–21].

Societal changes and the health care system’s organisation influence physicians’ roles and responsibilities. To learn how these changes impact moral distress over time, longitudinal studies are needed [22]. Few longitudinal studies of moral distress exist, and the available ones are of short duration (weeks or months) [23–25]. The Norwegian Physician Panel makes it possible to compare the same physicians over time.

Since 2004, the number of physicians in Norway has increased by 42% (from 18080 in 2004 to 29335 in 2021), and the fraction of female physicians increased from 35% to 53% (see Table 1). In this period, several new healthcare reforms and new health legislations have been implemented – such as increased patient rights. There have also been demographic shifts towards older patients, more medical services provided, and increased patient demands [26]. Although Norway was not hit as hard as many other countries by the COVID-19 pandemic, the pandemic significantly impacted the healthcare providers’ working conditions and workload in late 2020 – early 2021[19,27].

**Table 1.** Gender, age, and place of work among respondents (<□70) compared to physicians working in Norway (<□70) in 2021 and 2004.

|  | Responders in<br>2004 study<br>(n= 1004) | Physicians working<br>in Norway 2004<br>(n= 18080) | Responders in<br>2021 study<br>(n= 1639) | Physicians working<br>in Norway 2021<br>(n= 29335) |
| --- | --- | --- | --- | --- |
| Gender, proportion women | 32,9 % | 35,5 % | 52,8 % | 53,1 % |
| Average age, years | 49,1 | 45,1<sup>i</sup> | 46,7 | 44,1 |
| General practioners | 28,7 %<sup>ii</sup> | 23,6 % | 20,6 % | 22,8 % |
| Hopsital physicians | 54,1 %<sup>ii</sup> | 53,7 % | 63,5 % | 57,0 % |
| Others | 17,1 % | 21,2 % | 15,9 % | 20,2 % |

<sup>i</sup>2003 values are used since 2004 values were not reported
<sup>ii</sup> 2002 values are used since 2004 values were not reported

Our objective with this study was to explore and compare the physicians’ reported moral distress 17 years after the first study and look closer into factors that might explain their responses.

## Method

### Aim and design of the study

We examined how varying aspects of resource allocation, fairness concerns and acting against one’s conscience represented moral stress for physicians in Norway and how this changed over time. Comparisons were made between surveys of the Norwegian Physician Panel (NPP) in 2004 and 2021. We examined how age, gender, and professional position related to reported levels of moral distress.

### Participants

The NPP consists of a sample of physicians representing physicians working in Norway regarding age, gender, and workplace (hospital or primary care). Its representativity is measured against the Norwegian Medical Association’s register of members, where more than 90% of physicians in Norway are members. It was established in 1994 and has been complemented with new physicians regularly as existing members retire or withdraw for other reasons. The members are surveyed biennially through postal or digital questionnaires. The number of participants varies between 1500 and 2400, and the response rates are between 65% and 75%.

### Ethics

#### Patient and public involvement

Patients or the public were not involved in the design, conduct, reporting or dissemination of the results from this study. The reason was that the study was planned in 2003 -2004 when this requirement was not in place, and the research focused on moral distress among physicians.

#### Main outcome measures

##### Moral distress

The survey instrument was developed for the 2004 study, drawing upon Kälvemark’s empirical studies among healthcare providers in Sweden [18]. The same moral distress questions were used in 2004 and 2021. The introductory text was: «*Below, some situations are described which might lead to ethical dilemmas or “moral distress”. To what degree do you experience these kinds of situations as stressful?*” The statements are shown in Table 1. The participants scored the nine statements on a four-point Likert scale («Not distressing at all», «A little distressing», «Somewhat distressing», and «Very distressing»). In addition, there was a response alternative, «Don’t know» (2004)/«Not relevant for me» (2021).

##### Other variables

Gender was defined as male and female^1^, and age was reported as a continuous variable. We grouped the physicians according to workplace; general practitioners, hospital doctors, and others (laboratory medicine, academic or pure administrative work).

##### Analyses

The data were analysed with descriptive statistics (frequency distributions, crosstabs and chi square), in addition to multinomial logistic regression to identify the effect of the physician’s gender, age and workplace.

## Results

Response rates were 67% (1004/1499) in 2004 and 71% (1639/2316) in 2021. The samples are compared to working physicians in Norway in 2021 and 2004. Table 1 depicts the characteristics of the sample and the population of physicians.

### Moral distress

Table 2 shows the responses to the nine statements ranked from more to less morally distressing in 2021. The ranking had an almost similar pattern in 2004. There was a significant *increase* from 2004 to 2021 in reported moral distress for two statements, a significant *decrease* in four statements, and no significant changes in three statements.

**Table 2.**
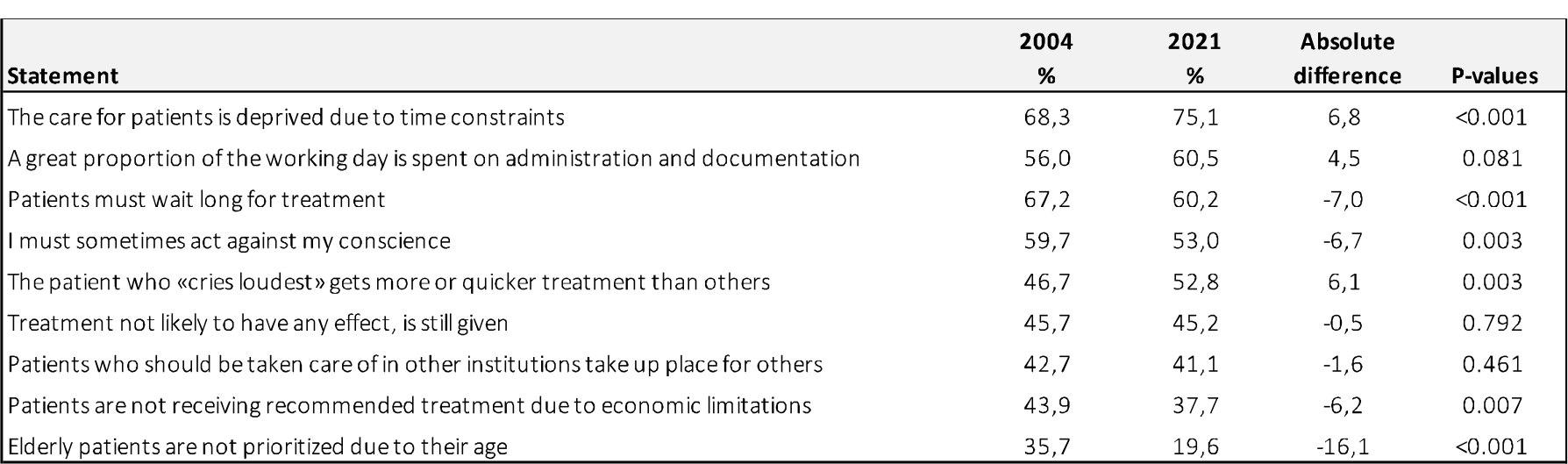
Physicians who responded: “Somewhat distressing” and “Very distressing” to the nine statements on moral distress shown in % in 2004 and 2021. Those who did not respond or responded “Don’t know” (2004) or “Not relevant for me” (2021) are excluded from the analysis.

| Statement | 2004<br>% | 2021<br>% | Absolute<br>difference | P-values |
| --- | --- | --- | --- | --- |
| The care for patients is deprived due to time constraints | 68,3 | 75,1 | 6,8 | <0.001 |
| A great proportion of the working day is spent on administration and documentation | 56,0 | 60,5 | 4,5 | 0.081 |
| Patients must wait long for treatment | 67,2 | 60,2 | -7,0 | <0.001 |
| I must sometimes act against my conscience | 59,7 | 53,0 | -6,7 | 0.003 |
| The patient who «cries loudest» gets more or quicker treatment than others | 46,7 | 52,8 | 6,1 | 0.003 |
| Treatment not likely to have any effect, is still given | 45,7 | 45,2 | -0,5 | 0.792 |
| Patients who should be taken care of in other institutions take up place for others | 42,7 | 41,1 | -1,6 | 0.461 |
| Patients are not receiving recommended treatment due to economic limitations | 43,9 | 37,7 | -6,2 | 0.007 |
| Elderly patients are not prioritized due to their age | 35,7 | 19,6 | -16,1 | <0.001 |

Complete distributions of responses, including “Don’t know” or “Not relevant for me”, are given in Supplement Table 1.

### Explanatory factors

#### Gender

Both in 2004 and 2021, women reported more moral distress in all statements. In 2021, the gender difference was significant in six of the nine statements when controlling for age and workplace: acting against one’s conscience, time constraints, prioritising those crying the loudest, waiting time, age discrimination, and economic limitations (p-values from <0.001 to 0.019). In 2004, women reported significantly more moral distress for “Elderly patients are not prioritised due to their age” (p-value 0.01).

The data from all regression analyses are presented in Supplement Table 2.

#### Age

In 2021, there was a significant age effect, when controlling for gender and workplace, on moral distress in six of the statements, such that lower age correlated with more moral destress. The statements were time constraints (P-value <0.001), patient crying the loudest getting more and faster treatment (p-value 0.001), time used on documentation and administration, providing treatment without effect, treatment in an unsuitable institution, and acting against one’s conscience (p-values 0.003 to 0.005). The pattern was the opposite on two statements: Patients not being prioritised due to higher age (both in 2021 and 2004) and, in 2004, providing treatment without effect. Here, the likelihood of reporting moral stress increased with age when controlling for gender and workplace.

#### General practitioners versus other clinicians

Both in 2004 and 2021, hospital physicians reported more moral distress due to economic limitations (p-values <0.001) and time constraints than the GPs. In 2021, hospital physicians reported more moral distress concerning patients being treated in an unsuitable institution (p-value 0.023), while in 2004, they reported more moral distress concerning the provision of treatment without effect (p-value 0.002). GPs reported significantly more moral distress than hospital physicians concerning providing more and faster treatment to those who cry the loudest (p-value 0.018) and time used on documentation and administration in 2021 (p-value 0.05).

## Discussion

Reported moral distress increased significantly from 2004 to 2021 for the statements concerning deprived care due to time constraints, and patients who cry the loudest get better and faster treatment than others. However, significantly less moral distress was reported in 2021 than in 2004 for the statements on long waiting times for treatment, not providing treatment due to economic limitations or the patient’s old age and acting against one’s conscience. Moral distress for seven of the statements was reported as high or somewhat high among more than 40% of the physicians in both years. Being a woman was a predictor of reporting higher moral distress related to most statements in 2021. The physicians’ age and position influenced the reported distress for some of the situations, where hospital physicians reported more distress related to time constraints and economic limitations, and young physicians tended to report more moral distress than older, the latter indicating the importance of supervision and ethics support for young doctors.

### More resources and more physicians, but less time for direct patient care?

Deprived care due to time constraints and time used on documentation and administration was reported as more distressful in 2021 than in 2004 and was the most distressful statement in both years. As many as 3/4 of all physicians found time constraints very or somewhat morally distressing in 2021. In this period, the health care budget increased substantially, and the number of physicians increased from 18080 to 29335. Yet, in the same period, the proportion of the physicians’ working hours seems to be pushed towards documentation, meetings, administrative tasks and follow-up of patients with more complex and time-consuming needs [28–30]. This can indicate that the organisation of the services and the physician’s time and tasks, more than lack of economic resources, are drivers for their moral distress. The change of work tasks has been one of the explicit reasons for general practitioners leaving their positions in recent years, causing a massive shortage of primary health care services [31]. Our study confirms that general practitioners report the most moral distress concerning the time used on documentation and administration.

Having sufficient time for their patients is highly valued by physicians [32]. A common complaint from patients is that physicians do not have enough time to listen or care for them, and medical students are taught communication techniques to hinder the patient from feeling the physician is speeding the consultation. Being confronted with a reality where their’ and the patients’ expectations can’t be fulfilled due to conflicting demands is stressful and affects role and self-identity. Studies show that being hindered from providing the care you believe the patient needs due to lack of time is a strong driver for experiencing moral distress [3].

### Decreased moral distress concerning economic limitations and waiting time

Moral distress due to long patient waiting times was high in both years. Interestingly, fewer physicians report high moral distress due to patients not receiving recommended treatment due to economic constraints and long waiting times in 2021 than in 2004. This might be due to more resources in the health care system, reforms regulating patient rights to access specialised health care, and special “rapid access”-referral procedures for patients with potentially severe conditions (such as cancer) [33]. It also confirms that moral distress due to time constraints is more closely linked to organisation and task-shifting than economic deprivation. Another interpretation could be that doctors resign themselves to a scarcity situation lasting over time and can experience that moral indignation or distress is of no avail. For example, in psychiatry, longer waiting times are documented[34]

### Fairness concerns at the bedside

There was a significant increase in reported moral distress with the statement that can indicate unfair distribution of treatment; “The patient who cries the loudest gets more and quicker treatment than others” from 2004 to 2021. This might imply that the 2021 patients (and next of kin) are more demanding and focused on their rights. Giving priority to the most trying patient is considered unfair but challenging to resist, and thus leads to moral distress [32,35].

In 2021 and 2004, almost half of our respondents found it very or somewhat distressing that futile treatment is still given. Studies show multiple and strong drivers for providing diagnostics and therapy without effect, and less experienced physicians are the ones who order the most unnecessary tests and are less likely to withhold or withdraw treatment [36,37]. Our study confirms this as the youngest physicians report the highest moral distress related to providing care without documented effect and to patients who cry the loudest. This might reflect their self-consciousness and internal struggle when acting against what they know is the fairest distribution.

### Decrease in moral distress related to age discrimination – reduced or perceived less problematic?

There was a significant decrease in the statement about age discrimination. One explanation is that there is less age discrimination in 2021 than in 2004. More treatments for diseases mostly hitting elderly patients have been implemented, and drugs previously only provided for patients below an age threshold are now available for all patients with the same condition. At the time we collected our data, it was just agreed that the elderly and the most vulnerable persons should be prioritised for the first COVID-19 vaccines (vaccination started at the end of December 2020), and this might have affected our responders.

Another explanation can be that physicians find that not treating patients due to their frailty and old age is morally defendable. In 2009, a national guideline for decision-making processes concerning withholding or withdrawal of care for severely sick and dying patients was developed and implemented [38]. Also, the latest national priority-setting legislation clarified that age is not a priority-setting criterion but that treatment for very old and sick patients might not be provided due to the reduced chance of effect of the therapy and lesser score on the severity of disease criteria [39]. It is hardly a surprise that older physicians find it morally distressing that elderly patients are not prioritised patients-their identification with patients can explain it.

### Decrease in moral distress related to acting against one’s conscience - a good or alarming sign?

Over half of the physicians reported moral distress related to acting against their conscience in both years. Still, the number decreased significantly from 2004 to 2021. One might expect that acting against what you believe is ethically correct will always be experienced as morally distressing. Therefore, 59.9% in 2004 and 53.0% in 2021 reported somewhat or very much moral distress is low. But experiencing moral activation due to challenged integrity - like acting against one’s conscience, does not necessarily lead to moral distress [5]. Tigard and others point out how moral activation reveals and affirms some of our most essential concerns as moral agents, and it can lead to moral maturation [40]. From this perspective, the decrease could be a good sign, indicating that the physicians have adjusted to their role, which involves making compromises and accepting their non-optimal reality. More explicit clinical guidelines and more explicitly regulated responsibility might also influence the level of moral distress connected to your ethical consciousness. Another explanation could be that being less activated by acting against one’s conscience is a symptom of high moral distress. Other studies show that there has been increased job stress and decreased life satisfaction among physicians, as well as alienation and despair among groups of physicians in this period [30,41]. Also, in 2021, there was a significantly higher response among younger physicians than in 2004. While it is well known that younger physicians report more moral distress than their more experienced colleagues [19,42], our findings indicate that other factors are in play, not age alone. More research is needed to explore this further.

### Women report more moral distress than men

More women reported moral distress both in 2004 and 2021. Other studies also show gender-mediated behavioural differences that might impact the difference in reported moral distress among physicians. [43]. Female physicians are more likely to spend more time on patient communication and electronic medical journal work, show higher empathy behaviour, provide patient-centred care, and are less comfortable in decision-making under uncertainty [44–46]. These behavioural differences might be due to cultural expectations of women and gender norms in health care. Why the gender differences are more significant in our 2021 study than in the 2004 study needs further attention.

### Way forward

In the early summer of 2023 in Norway, a post on social media described how stressful work conditions led a young mother and physician to commit suicide. The post initiated a massive response from physicians, primarily women, many expressing their vulnerability, fear, frustrations, and system factors hindering working conditions aligning with a good life [47]. The peer-to-peer physician services and the special health care services for physicians in Norway have experienced a steep increase in requests for support, particularly from young women [48]. Also, studies and reports from other countries report physician resignations and despair related to their work conditions [49,50]. These alarming facts should lead to further investigations and possible measures.

There is a need for discussions on improvements in the system, leadership responsibility and prioritisation of what physicians should spend their time on. We must better prepare our medical students and support our physicians to handle issues like time scarcity, demanding patients, and withholding or withdrawing futile care in ways that are fair and can align with their self-perception of being good physicians. A Norwegian Whitepaper on human resources in health care concluded that the ongoing and future lack of health providers can’t be solved only by employing more providers and that the health care system needs to adjust to higher demands [51]. Emerging research on strategies to increase resilience and robustness among healthcare providers shows promising results on how moral distress can decrease among staff, prohibit resignation and improve work satisfaction even in an ongoing pandemic situation [52]. These strategies can include helping staff recognise moral distress responses and various coping mechanisms, thus increasing their ethical competency and confidence in dealing with complex situations (17). Our study shows this is particularly important for young and less experienced physicians. Our results can bring valuable insight into these discussions, providing solid empirical evidence on what situations impose moral distress on physicians.

### Strengths and limitations

Our data was collected in December 2021-January 2021, a “quiet” period in the pandemic in Norway. Still, the COVID-19 pandemic led to many new tasks and challenges in all parts of the healthcare system. We only found an increase in reported moral distress concerning time constraints and demanding patients receiving treatment faster and better than others in 2021 compared to 2004. This increase might be pandemic-dependent, but the no change or decrease in the other statements is hard to explain with the pandemic. Numerous papers have been published on healthcare providers’ increased moral distress during the COVID-19 pandemic. But few of them are from a representative sample of the population, and none, as far as we know, have compared it to data collected at different points in time. Our heterogeneity in responses regarding year and gender, age and specialisation should lead to more curiosity on the pandemic effect on experienced moral distress in other data materials.

Our study’s relatively high response rate (70%) is a strength. This is higher than for other surveys of the medical profession [22]. The sample also represents the population of practising physicians in Norway in key aspects like gender, age and workplace [23]. This provides a reasonable basis for generalisation but does not entirely rule out the possibility of nonresponse bias.

## Conclusion

Our study is one among very few which shows how morally distressing physicians experience resource scarcity, overtreatment, and unfair distribution over 17 years. Clinicians are among our most precious healthcare resources, and we need more knowledge on how to ensure they can act according to ethical standards and keep their integrity in the times coming with increased demands and fewer resources available. A particular focus should be on understanding and working on the high moral distress among young women to make them thrive in the profession and not get burned out.

## Data Availability

All data produced in the present study are available upon reasonable request to the authors.

---

**Supplement table 1.**
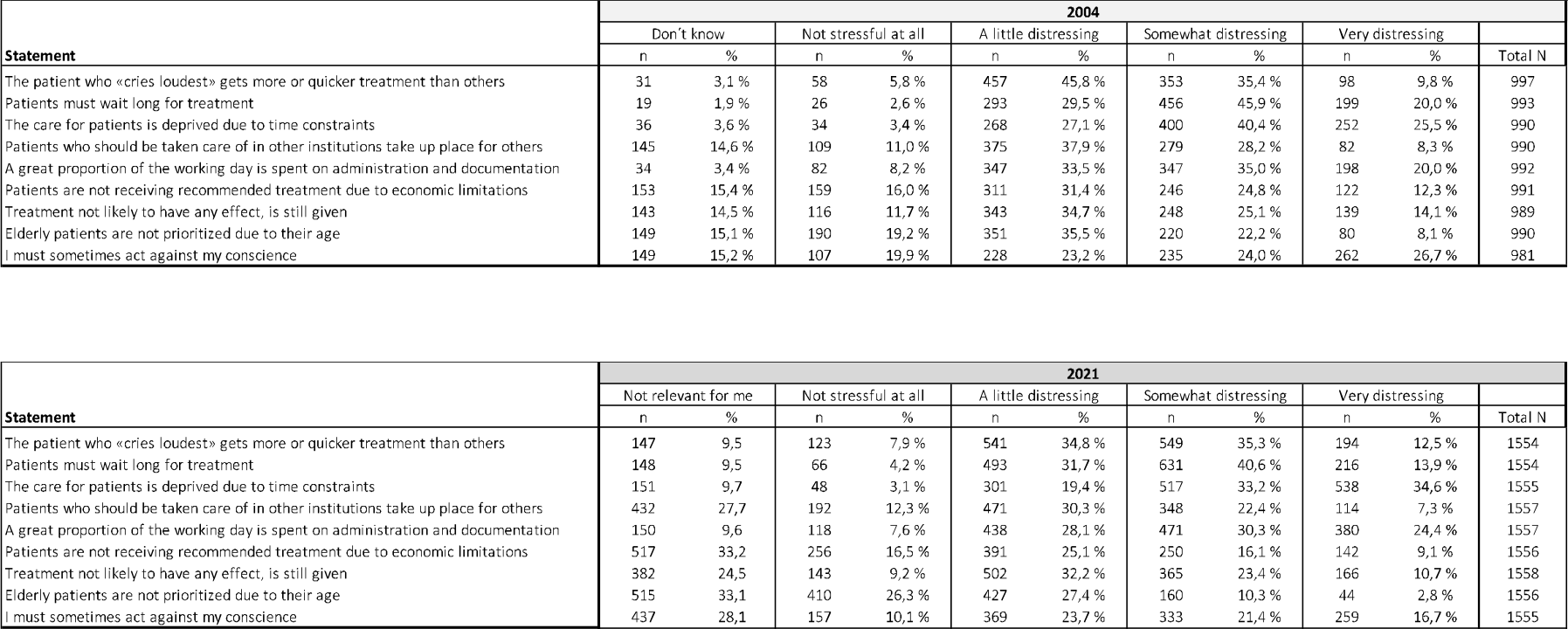
Responses to the nine statements in 2004 and 2021 shown in numbers and %.

| Independent variables | B | SE | sig | Exp(B) |
| --- | --- | --- | --- | --- |
| Age | -.003 | .009 | .733 | .997 |
| Sex | .282 | .183 | .123 | 1.326 |
| Workplace | .300 | .171 | .079 | 1.350 |
| Constant | -.131 | .462 | .776 | .877 |

| Independent variables | B | SE | sig | Exp(B) |
| --- | --- | --- | --- | --- |
| Age | .007 | .009 | .472 | 1.007 |
| Sex | .222 | .198 | .262 | 1.249 |
| Workplace | .053 | .182 | .772 | 1.054 |
| Constant | .319 | .491 | .516 | 1.376 |

| Independent variables | B | SE | sig | Exp(B) |
| --- | --- | --- | --- | --- |
| Age | -.012 | .010 | .246 | .997 |
| Sex | .240 | .206 | .244 | 1.326 |
| Workplace | -.899 | .182 | **<.001** | .407 |
| Constant | 1.604 | .519 | .002 | 4.970 |

| Independent variables | B | SE | sig | Exp(B) |
| --- | --- | --- | --- | --- |
| Age | -.011 | .009 | .733 | .997 |
| Sex | .282 | .183 | .123 | 1.326 |
| Workplace | .300 | .171 | .079 | 1.350 |
| Constant | -.131 | .462 | .776 | .877 |

| Independent variables | B | SE | sig | Exp(B) |
| --- | --- | --- | --- | --- |
| Age | .008 | .009 | .406 | 1.008 |
| Sex | -.044 | .186 | .812 | .957 |
| Workplace | -.237 | .172 | .169 | .789 |
| Constant | .075 | .471 | .873 | 1.078 |

| Independent variables | B | SE | sig | Exp(B) |
| --- | --- | --- | --- | --- |
| Age | .016 | .010 | .109 | 1.016 |
| Sex | .323 | .206 | .117 | 1.381 |
| Workplace | -.678 | .193 | **<.001** | .508 |
| Constant | -.984 | .518 | .057 | .374 |

| Independent variables | B | SE | sig | Exp(B) |
| --- | --- | --- | --- | --- |
| Age | .020 | .010 | .037 | 1.020 |
| Sex | -.188 | .203 | .355 | .829 |
| Workplace | -.569 | .188 | .002 | .566 |
| Constant | -.979 | .500 | .051 | .376 |

| Independent variables | B | SE | sig | Exp(B) |
| --- | --- | --- | --- | --- |
| Age | .038 | .010 | **<.001** | 1.038 |
| Sex | .556 | .215 | .010 | 1.744 |
| Workplace | .036 | .193 | .853 | 1.036 |
| Constant | -2.750 | .557 | **<.001** | .064 |

| Independent variables | B | SE | sig | Exp(B) |
| --- | --- | --- | --- | --- |
| Age | .005 | .010 | .629 | 1.005 |
| Sex | .371 | .202 | .067 | 1.449 |
| Workplace | -.188 | .183 | .304 | .829 |
| Constant | .114 | .507 | .822 | 1.121 |

Sex: Male = 0 Female = 1
Workplace: Hospital = 0 GP = 1

| Independent variables | B | SE | sig | Exp(B) |
| --- | --- | --- | --- | --- |
| Age | -.017 | .005 | .001 | .983 |
| Sex | .360 | .125 | .004 | 1.433 |
| Workplace | .332 | .141 | .018 | 1.394 |
| Constant | .572 | .262 | .030 | 1.796 |

| Independent variables | B | SE | sig | Exp(B) |
| --- | --- | --- | --- | --- |
| Age | -.001 | .005 | .887 | .999 |
| Sex | .339 | .126 | .007 | 1.404 |
| Workplace | .173 | .142 | .222 | 1.189 |
| Constant | .172 | .265 | .516 | 1.188 |

| Independent variables | B | SE | sig | Exp(B) |
| --- | --- | --- | --- | --- |
| Age | -.027 | .006 | <.001 | .974 |
| Sex | .450 | .145 | .002 | 1.568 |
| Workplace | -.427 | .156 | .006 | .653 |
| Constant | 2.176 | .306 | <.001 | 8.810 |

| Independent variables | B | SE | sig | Exp(B) |
| --- | --- | --- | --- | --- |
| Age | -.014 | .006 | .026 | .986 |
| Sex | -.096 | .140 | .493 | .908 |
| Workplace | -.383 | .168 | .023 | .682 |
| Constant | .385 | .298 | .197 | 1.469 |

| Independent variables | B | SE | sig | Exp(B) |
| --- | --- | --- | --- | --- |
| Age | -.016 | .005 | .003 | .984 |
| Sex | .085 | .127 | .507 | 1.088 |
| Workplace | .284 | .145 | .051 | 1.328 |
| Constant | 1.065 | .269 | <.001 | 2.902 |

| Independent variables | B | SE | sig | Exp(B) |
| --- | --- | --- | --- | --- |
| Age | .003 | .006 | .606 | 1.003 |
| Sex | .351 | .149 | .019 | 1.421 |
| Workplace | -.619 | .168 | <.001 | .538 |
| Constant | -.656 | .315 | .037 | .519 |

| Independent variables | B | SE | sig | Exp(B) |
| --- | --- | --- | --- | --- |
| Age | -.016 | .006 | .007 | .984 |
| Sex | .058 | .136 | .668 | 1.060 |
| Workplace | -.051 | .151 | .735 | .950 |
| Constant | .375 | .293 | .201 | 1.455 |

| Independent variables | B | SE | sig | Exp(B) |
| --- | --- | --- | --- | --- |
| Age | .034 | .008 | <.001 | 1.034 |
| Sex | .470 | .196 | .016 | 1.600 |
| Workplace | .336 | .200 | .092 | 1.400 |
| Constant | -3.396 | .414 | <.001 | .034 |

| Independent variables | B | SE | sig | Exp(B) |
| --- | --- | --- | --- | --- |
| Age | -.017 | .006 | .005 | .983 |
| Sex | .495 | .140 | <.001 | 1.641 |
| Workplace | .043 | .151 | .775 | 1.044 |
| Constant | .497 | .298 | .095 | 1.644 |

## Footnotes

1 In 2021, we also gave the option to type of “other gender identity”. Two ticked off this, and in the further analysis, one was moved to male and one to female as the numbers were too low to do separate studies on

